# Women’s perceptions regarding access to pelvic health physiotherapy: A qualitative study

**DOI:** 10.1101/2025.09.09.25335426

**Authors:** Valérie Boulanger, Mélanie Morin, Stéphanie Bernard, Anne Hudon, Laurence Chevalier, Kassandra Gamache, Rose-Marie Bourget, Camille Simard, Olivia Fortier, Ariane Gendron, Allyson Pépin, Geneviève Morin, Isabelle Girard, Annie-Kim Gareau-Labelle, Valérie Charbonneau, Ève Levasseur, Gabrielle Déry-Rouleau, Kadija Perreault

**Affiliations:** Center for Interdisciplinary Research in Rehabilitation and Social Integration, Centre intégré universitaire de santé et de services sociaux de la Capitale-Nationale, Quebec City; School of Rehabilitation Sciences, Faculty of Medicine, Université Laval, Quebec City; Research Center of the Centre Hospitalier Universitaire de Sherbrooke, Sherbrooke; School of Rehabilitation, Faculty of Medicine and Health Sciences, Université de Sherbrooke, Sherbrooke; Centre for interdisciplinary Research in Rehabilitation of Greater Montreal (CRIR), Montreal; School of Rehabilitation, Université de Montréal, Montreal; Department of Health Sciences, Université du Québec à Chicoutimi, Chicoutimi; Centre hospitalier de l’Université de Montréal (CHUM), Montreal; Centre hospitalier de l’Université Laval (CHUL), Quebec City; Groupe de médecine familiale universitaire (GMF-U) du Sud de Lanaudière, Lanaudière; Centre intégré de santé et de services sociaux (CISSS) of Lanaudière, Lanaudière; Centre Hospitalier Universitaire de Sherbrooke, Sherbrooke

**Keywords:** pelvic health physiotherapy, access, women’s health, qualitative research, pelvic floor

## Abstract

**Purpose:** Pelvic floor dysfunctions (PDFs) are highly prevalent in adult women. Although pelvic health physiotherapy (PHP) has been found to provide evidence-based interventions to women presenting these conditions, access to such services may be limited. The objective of this study was to explore women’s perceptions and experiences regarding access to pelvic health physiotherapy (PHP).

**Methods:** This study was a qualitative descriptive study using semi-structured interviews. An interview guide was used to interview women having experienced PFD, who had or not accessed PHP services. The content of the interviews was transcribed and analyzed through inductive reflective thematic analysis. The transcripts were coded, and key themes were identified using an iterative process.

**Results:** The participants were 15 women over the age of 18 years. Three main themes were identified: 1) lacking guidance on PFDs and PHP services; 2) neglecting PFDs and their effects; 3) demanding access to quality PHP services.

**Conclusion:** The results of this study provide valuable insights into the access issues faced by women regarding their pelvic health. Findings highlight numerous unmet needs, among which the need to improve knowledge and awareness of PFDs, to address the trivialization of PFD symptoms, and to ensure access to PHP in rural areas.

## INTRODUCTION

Pelvic floor dysfunctions (PFDs) are highly prevalent among women, with 67.5% experiencing some form of PFDs during their lifetime (Kepenekci et al., 2011). Urinary incontinence and chronic pelvic pain are two of the most common PFDs, with prevalence rates ranging from 25% to 45% for urinary incontinence and 4% to 16% for chronic pelvic pain (Dydyk and Gupta, 2024; Leslie, Tran, and Puckett, 2024). PFDs have a significant impact on the physical, mental and sexual health (Hermansen, O’Connell, and Gaskin, 2010). For example, in a study by Hawke et al. (2022), women with chronic pelvic pain reported that it impacted intimate relationships, fertility, parenting, and work (Hawkey et al., 2022). PFDs occurring in the perinatal period can also have significant psychological impacts, having been associated with positive screening for post-partum depression (Gallego-Gómez et al., 2024; Till, As-Sanie, and Schrepf, 2019). In addition, given the high prevalence of PFDs and their widespread impact on the biopsychosocial well-being of individuals, these conditions present with a significant economic burden. In Canada, the combined direct and indirect costs associated with urinary incontinence alone were estimated to be between 2.6 and 8.5 billion dollars annually (INESSS, 2022).

Evidence strongly supports pelvic health physiotherapy (PHP) as the first-line treatments for several PFDs, including urinary incontinence, pelvic organ prolapse, and chronic pelvic pain (Allaire et al., 2024; Cardozo et al., 2023; Tunn, Baessler, Knüpfer, and Hampel, 2023). PHP involves a combination of several interventions such as education, myofascial techniques, dilatation and pelvic floor muscle training (Boyle, Hay-Smith, Cody, and Mørkved, 2014). Despite the evidence supporting the effectiveness of multimodal physiotherapy interventions in treating PFDs, it has been suggested that access to PHP services may be challenging for women (Boyle, Hay-Smith, Cody, and Mørkved, 2014). Certain barriers that have been identified include financial and time constraints (Zoorob et al., 2017). Based on Lévesque et al.’s (2013) framework on access to healthcare services, several factors can hinder an individual’s access to healthcare services and optimal healthcare utilization. Indeed, access results from the individual’s capacity to perceive, seek, reach, pay and engage in care, in interaction with the approachability, acceptability, availability, affordability and appropriateness of services provided by healthcare providers, institutions and systems. Little research has nonetheless been conducted to understand the challenges faced by women regarding accessing to PHP services. Therefore, the objective of this study was to explore the perceptions and experiences of women regarding access to PHP services.

## MATERIALS AND METHODS

### Design and study population

A qualitative descriptive design was used. This approach is particularly effective for exploring perceptions and emotions, providing rich insights into the lived experiences of individuals (Busetto, Wick, and Gumbinger, 2020). This study is the second phase of a larger research project on access to PHP in Quebec, Canada’s second most densely populated province, using a mixed sequential explanatory design (Plano Clark, 2019). The first phase involved two cross-sectional electronic surveys targeting women 18 or over who had experienced chronic pelvic pain, including vulvar pain, for at least three months, as well as women having given birth at least once, with the most recent delivery occurring within the previous four years. The women also had to live in Quebec, where PHP services are primarily offered in private settings (Deslauriers et al., 2017). Building on the findings from this initial phase, the current study was meant to further examine women’s experiences and perceptions of access to PHP services through their own narratives using semi-structured interviews.

Participants were selected among women who had completed at least one of the surveys in the first phase. Women who indicated in the survey being interested in participating in an interview were contacted via email to answer questions designed to clarify their profiles. This also allowed to promote diversity within the interview participants in order to capture a broad range of experiences and perspectives (e.g. age, ethnicity, location, having accessed PHP services or not) (Colorafi and Evans, 2016). All participants provided informed consent to take part in an interview.

The study was approved by the Ethical Review Board of the *Centre intégré universitaire de santé et de services sociaux de l’Estrie-Centre hospitalier universitaire de Sherbrooke* (MP-31-2022-3987).

### Data collection

Interviews were conducted and recorded securely via Microsoft Teams or Zoom. An interview guide was developed based on the research objectives, prior studies on access to services, and preliminary findings from the first phase of the study. The guide was also developed with the involvement of three patient partners. This allowed to refine the content through an iterative process to enhance clarity, reorganize content, and modify questions as necessary (DeJonckheere and Vaughn, 2019). The two interviewers (KG and RMB) received training and mentorship from team members experienced in qualitative methods. Two interviews were first conducted to validate the content of the interview guide and procedures (Berre and Dumoulin, 2024). Feedback and discussions between team members (KG, RMB, LC, KP, MM, SB) based on the recordings of the first two interviews led to further refinements of the interview guide. Participants also completed a socio-demographic questionnaire to help characterize sample.

### Data analysis

Audio-recordings of all interviews were first transcribed verbatim by a professional transcriber. The transcripts were then reviewed by a member of the research team to ensure completeness, accuracy, and the anonymity of individuals or places mentioned in the interviews (Baker et al., 2016). The interviews were then imported into QDA Miner Lite (version 3.0) to facilitate analyses.

An inductive reflective thematic analysis approach was used to identify themes from the women’s narratives, reflecting their perceptions and lived experiences (Braun and Clarke, 2019). Thorough familiarization with the data was achieved through multiple readings of the transcripts, enhancing the quality of the analysis. Each interview was coded by one member of the research team (LC or VB) and subsequently analysed by another member. This pairwise analysis facilitated the identification of major codes and the development of sub-codes. The analysis took into account not only the women’s words but also non-verbal elements such as pauses and laughter, which were carefully considered during the analyses process (Durkin, Jackson, and Usher, 2020). Codes were systematically grouped and refined to eliminate redundancies and improve the clarity of the analysis. Following the completion of the code tree led by the first author, the team collaboratively and iteratively identified the key themes and subthemes that best represented the main findings of the study, in alignment with its objective to explore women’s experiences and perceptions of access to PHP. Prominent quotes reflecting these core themes and subthemes were selected to ensure that the women’s voices were authentically represented in the findings.

## RESULTS

### Description of participants

Interviews were conducted with 15 women. They lasted between 37 and 62 minutes. The average age of the participants was 39 years (± 15) and they mostly reported living in urban (86.7%) areas. Over 55% of participants had a household income of $120,000 or more. More than half of the women (60.0%) had given birth in the previous four years or had experienced a PFD following childbirth or pregnancy. Additionally, 73.3% had experienced pelvic symptoms or chronic pelvic pain for over three months. The sample included both women who had not consulted for PHP (40%) and those who had (60%), in private or public settings or both. Characteristics of participants are presented in Table 1.

**Table 1.** Characteristics of participants (n=15)

| Characteristics |  | *n* (%) |
| --- | --- | --- |
| Age (years) | < 30 | 4 (26.7) |
|  | 31-50 | 8 (53.3) |
|  | > 51 | 3 (20.0) |
| Marital status | Single | 3 (20.0) |
|  | Living with partner | 7 (46.7) |
|  | Married | 5 (33.3) |
| Born in Quebec | Yes | 13 (86.7) |
|  | No | 2 (13.3) |
| Household income (\$ CAD) | < 40 000 | 3 (20.0) |
|  | 40 000 à < 80 000 | 1 (6.7) |
|  | 80 000 à < 100 000 | 1 (6.7) |
|  | 100 000 à < 120 000 | 2 (13.3) |
|  | ≥ 120 000 | 8 (53.3) |
| Education level | High school | 2 (13.3) |
|  | College | 3 (20.0) |
|  | Bachelor's degree | 4 (26.7) |
|  | Graduate degree | 6 (40.0) |
| Employment status | Part-time | 2 (13.3) |
|  | Full-time | 8 (53.3) |
|  | Retired | 2 (13.3) |
|  | Student | 2 (13.3) |
|  | Maternity leave | 1 (6.7) |
| Place of residence | Rural | 2 (13.3) |
|  | Urban | 13 (86.7) |
| PHP consultation | Did not consult | 6 (40.0) |
|  | Consulted in public setting | 2 (13.3) |
|  | Consulted in private setting | 5 (33.3) |
|  | Consulted in both public and private | 2 (13.3) |
| Women who gave birth in the previous 4 years |  | 9 (60.0) |
| Women who had experienced pelvic pain for more than 3 months |  | 11 (73.3) |

### Perceptions and experiences regarding access to PHP

Women’s perceptions and experiences regarding access to PHP were found to fall under three themes: 1) lacking guidance on PFDs and PHP services, 2) neglecting PFDs and their effects, 3) needing access to quality PHP services.

#### Theme 1: Lacking guidance on PFDs and PHP services

The first theme expresses how participants felt they lacked guidance regarding their condition and the services available to them. This need for enhanced guidance was related to four main issues, reported here as sub-themes: knowledge gaps surrounding PFDs among healthcare professionals, knowledge gaps surrounding PFD management, lack of awareness among women about PFDs, and hard-to-access information.

Many women detailed how health professionals lacked knowledge on PFDs, which affected their ability to refer patients to the most appropriate treatment options. Some healthcare professionals didn’t seem to know what symptoms to look for and how these issues can profoundly impact one’s life. For example, one participant mentioned needing to consult multiple health care professionals for her PFD, stating:

> *“I experienced perineal issues that led me to consult five different healthcare professionals… I felt very poorly equipped, very poorly supported in dealing with this issue.” (P7, age 25-34, chronic pelvic pain and post-partum, consulted in private setting)*

Another challenge experienced by participants was that even when healthcare professionals were able to identify a PFD, they often lacked knowledge on how to manage it, or how PHP could benefit patients. This resulted in frustration, as described by one participant:

> *“For him, well, he’s a urological surgeon, […] he performed a cystoscopy to check if everything was fine, and he said, ‘This is it, madam, it’s interstitial cystitis, and there’s nothing to be done, but you can try Elmiron.’ I was so outraged! When I started discovering these options that were available [PHP], I mean, it’s unbelievable that he didn’t know about this!” (P13, age 65-74, chronic pelvic pain, consulted in public setting)*

This situation was echoed by another participant, whose family physician seemed unaware of how PHP could help women with various PFDs. When asked if her doctor was aware of her urinary leakage, she responded:

> *“Yes, my family doctor is aware […] Apparently, there’s nothing that can be done.” (P10, age 25-34, chronic pelvic pain and post-partum, did not consult)*

The lack of knowledge regarding pelvic health and PFDs also extended to women themselves. Indeed, many participants reported lacking knowledge to differentiate between what constituted a normal experience and what signaled an issue that should be addressed, as articulated by one participant:

> *“If there had been something that would have made me seek help. In fact, I think that not knowing—when we don’t even know it’s a muscle, or we don’t even know this part of our body—means that even if we have problems, we won’t seek help.” (P9, age 35-44, chronic pelvic pain and post-partum, did not consult)*

Participants mentioned that because information on PFDs is limited and not readily accessible, this hinders their ability to recognize symptoms by themselves and seek appropriate care. As one participant stated:

> *“We don’t advertise this [PHP], why don’t we talk about it? These monthly appointments we have with doctors to check in on how things are going—why isn’t there a pamphlet there? Why don’t we discuss it during those moments? There are videos on absolutely horrible things, but there’s absolutely nothing on pelvic floor physiotherapy, so why not!” (P14, age 35-44, chronic pelvic pain and post-partum, consulted in public setting).*

Participants expressed that healthcare professionals and organizations should advocate for better access to care for women which would allow to empower them. As one woman said:

> *“We can’t carry the burden of something we don’t know about (nervous laughs). So, we’re stuck in a vicious cycle.” (P9, age 35-44, chronic pelvic pain and post-partum, did not consult)*

Overall, these narratives highlighted a lack of knowledge about the benefits of PHP or about PFDs and were a clear call to be better guided.

#### Theme 2: Neglecting PFDs and their effects on women

The second theme lies on the importance of PFDs as a critical component of women’s overall health. Participants felt this was neglected. This neglect arose from a variety of factors, including the trivialization of symptoms by healthcare professionals, the normalization of symptoms by women themselves, conflicting sociocultural influences, and stigma associated with the pelvic floor.

Many women shared similar experiences associated with the trivialization of their symptoms during encounters with healthcare providers, this going beyond the lack of knowledge highlighted in the first theme. Some were told that their symptoms were normal, while others felt their concerns were dismissed and they were advised to continue living their lives as if the symptoms were of no significance. One participant said:

> *“Basically, she [the doctor] didn’t do much. You know, she didn’t investigate, she didn’t look into what was going on [regarding my urinary incontinence], but she told me that at the stage I was at, if I felt okay, I could resume physical activity.” (P4, age 25-34, post-partum, did not consult).*

This trivialization of symptoms paved the way to a normalization of symptoms by women themselves, which led them to refrain from seeking further services. Indeed, many participants expressed that when the main source of health information is the healthcare professional, you trust and believe them when they say the problem is normal. One woman stated:

> *“When I asked questions, when… I asked my doctor, […], it was always ‘Oh, it’s normal, it’s normal, it’s normal,’ so it’s like having discomfort is just normalized.” (P9, age 35-44, chronic pelvic pain and post-partum, did not consult)*

Participants also valued the opinions and recommendations made by people in their close environment. They turned to their close friends and family members for guidance, learning from their life experiences and them as models. Women mentioned that if they are told by either a medical professional or their close circle that their problem is normal, they might endure it and come to see it as normal themself. This, in turn, can lead other women to adopt the same perspective, as described by this participant:

> *“In fact, the majority of the moms [I know] didn’t seek consultation after that [giving birth]. Uh… and you know, for them, it’s not necessarily a very disabling situation either. It’s, it’s… you know, it’s sporadic, so they almost consider it normal to have a small incontinence problem occasionally.” (P15, age 35-44, chronic pelvic pain, did not consult)*

Open discussion about PFDs was also limited by the stigma associated with the pelvic floor, which was frequently associated with negativity and shame. Participants suggested that one way of breaking this taboo would be to facilitate conversations on these issues. As stated by one participant:

> *“I think that if it is talked about more and women realize that it’s a reality shared by many, it will help reduce the stigma and the taboo surrounding it. It would also support the idea of a campaign to emphasize the importance of discussing these issues with those around you to gain support.” (P5, age 25-34, chronic pelvic pain, did not consult)*

These statements point to the need to recognize PFDs and their impacts for women.

#### Theme 3: Needing access to quality PHP services

The third theme highlights the difficulties experienced by women in accessing and obtaining quality services. Participants who received services talked about the benefits of accessing PHP, including improvements in quality of life and the alleviation of symptoms. However, participants also identified several factors that negatively impacted access to quality PHP services like high costs of care in private settings, delays in accessing PHP services in public settings, restricted access to services and expertise in rural areas, and inconsistencies in the PHP services.

A frequently cited barrier to accessing PHP was affordability, with many women highlighting the high costs associated with having to turn to private settings to obtain services, due to limited service offers in public settings. These financial burdens could prevent women from seeking treatment or cause them to discontinue care prematurely. In addition, without publicly funded services or insurance coverage, many women would not have been able to receive treatments at all:

> *“I didn’t have to pay; it was covered […] I’m not sure I would have gone for five months if I had to pay $100 each time I went, because it’s even more expensive now—the last time I checked, it was up to $140. I can’t afford that.” (P13, age 65-74, chronic pelvic pain, consulted in public setting)*

Some women even stated that the high fees in private clinics led them to question if they really needed the services, because they had to choose between accessing this healthcare treatment and meeting their basic needs; hence possibly overlooking pelvic health as a need itself.

In addition, while PHP services can be accessed through both private and public healthcare systems in the local healthcare system, the possibility of consulting a pelvic health physiotherapist in the public sector was perceived as something only available to women if they had a practitioner who was aware of the existence of public PHP services, if this person was willing to provide a referral, and if the service was available in the region where they lived. As mentioned by this participant who expressed frustration with lengthy waiting times in a public setting:

> *“You know, the doctor… They’re the best person to assess it, but what’s the point of the doctor making a referral when there’s no access in the public system? So, [what if] the patient works at a convenience store, wants to get treated but can’t afford it, and there’s a huge waiting list in the public system? -what is she supposed to do!” (P3, age 25-34, chronic pelvic pain and post-partum, consulted in private setting)*

Long wait times and other barriers to accessing PHP were also reported to be problematic in both public and private settings, especially in remote areas:

> *“I think that living in a remote area is an added challenge because, you know, just by searching quickly […] if I find someone who offers [PHP services], that’s great—and that’s if they’re still practicing. So, you know, after that, it’s understood that the wait times to meet this person increase […] But it means that, well, living in a remote area becomes a disadvantage for my feminine health, you know.” (P15, age 35-44, chronic pelvic pain, did not consult).*

Even if they had the possibility to obtain services, a few women also reported an additional burden related to experiencing services that were not all of equal quality. One participant shared how she consulted two different physiotherapists for the same condition, but received services of different qualities according to her:

> *“The first physiotherapist I went to [for PHP services] […] she would just check me for like two minutes, then hand me some papers, and that was it. I was like, ‘Ok no, this isn’t working.’ The other one, though, spent maybe 45 minutes with me, sometimes even an hour! We would talk a lot at the beginning […] she asked me a lot of questions about how it [chronic pelvic pain] affected me psychologically, which I thought was great. She was amazing, really! The service she provided was super thorough…” (P7, age 25-34, chronic pelvic pain and post-partum, consulted in private setting)*

Figure 1 visually represents this latter theme as well as other themes and corresponding subthemes.

**Figure 1.**
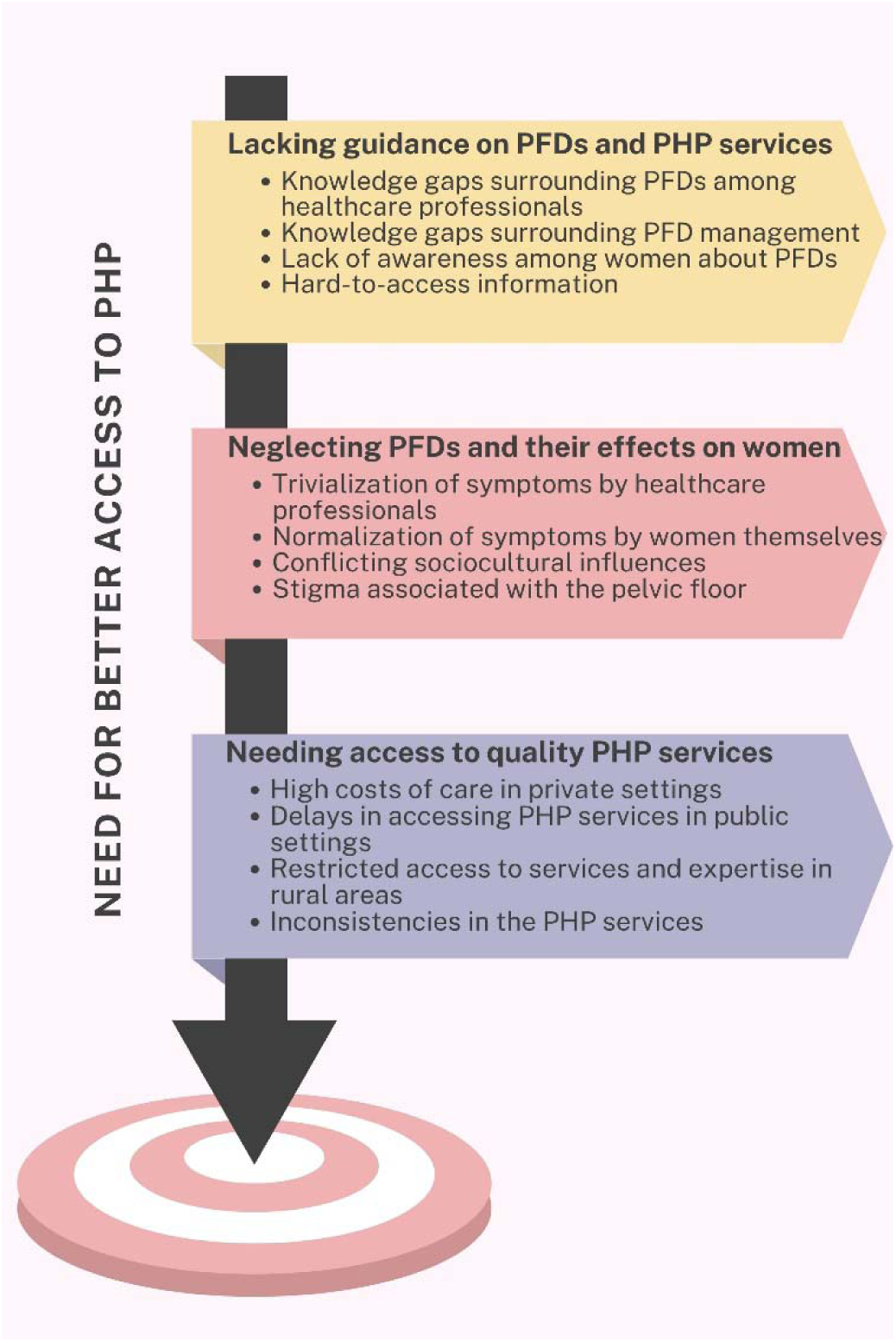
Representation of themes and sub-themes

## DISCUSSION

The aim of the study was to explore women’s perceptions and experiences regarding access to PHP services, regardless of whether or not they had consulted. Findings from this study reveal that numerous barriers hinder access to PHP services, even though they are recommended as a first-line approach for treating PFDs (Allaire et al., 2024; Cardozo et al., 2023; Tunn, Baessler, Knüpfer, and Hampel, 2023). The main themes identified in this study all highlight unmet needs in accessing PHP services: 1) lacking guidance on PFDs and PHP services, 2) neglecting PFDs and their effects on women, and 3) needing access to quality PHP services. These themes and associated subthemes relate to numerous dimensions of Levesque et al.’s conceptual framework of access to healthcare, highlighting gaps in both the characteristics of healthcare services (their accessibility)—such as approachability, acceptability, availability, affordability, and appropriateness—, as well as women’s corresponding abilities, that is the ability to perceive their need for care, to actively seek it, to reach, to afford, and to engage with the services (Levesque, Harris, and Russell, 2013). This framework was explicitly used in another study of accessibility of PHP (Green et al., 2024).

The first theme that was identified from the interviews was the significant lack of knowledge about key aspects of pelvic health. This gap in understanding, shared by both healthcare professionals and women, extends to symptoms associated with PFDs and the PHP services for the management of their condition. This results in an unrecognition of the potential for improving one’s condition and any possible benefits that may result from accessing and utilizing healthcare services by women, regardless of the severity of the symptoms experienced. Indeed, recognizing that a healthcare need is present is a crucial step to enable to seek and access care for that need (Levesque, Harris, and Russell, 2013). In the case of PFDs, this recognition requires some understanding of what a healthy pelvic floor is and which symptoms to be aware of. The findings of this study highlight that both women themselves and health professionals appear to overlook these needs because of a lack of knowledge on these issues and how to best manage PFDs. This is in line with the findings of a systematic review conducted by Fante et al. on women’s knowledge about pelvic floor dysfunctions, which also showed a lack of knowledge about PFDs among women, as well as the urgent need for improving education and provide better guidance on these dysfunctions and PHP services (Fante et al., 2019).

The second theme emphasizes what can be identified as another critical, yet related, barrier to accessing PHP services: neglecting PFDs and their impacts on women, including their physical health and well-being. Indeed, the results of this study show that participants often experienced being dismissed or that their symptoms were not taken into consideration, when accessing healthcare. Women feeling powerless or disregarded when it comes to their health is not a new phenomenon, as shown for instance in a 2007 study which concluded that women were often being nullified following gynecological consultations (Swahnberg, Thapar-Björkert, and Berterö, 2007). Participants further described that the trivialization of their symptoms by healthcare professionals led to women normalizing their own symptoms. Over time, this normalization can become entrenched across different generations or social circles, as heard from participants in this study. This may also further discourage open discussion and reinforce the stigma surrounding pelvic floor health(Jouanny, Abhyankar, and Maxwell, 2024). These interrelated elements appear to create a vicious cycle that could be broken by a better recognition of the impacts of PFDs on women’s health and creating contexts to facilitate discussion surrounding women’s needs.

The last theme raised by this study was needing access to quality PHP services. Participants expressed a recurring concern of high costs of these services pointing out that individuals with higher household incomes often have better insurance coverage. This allows them to access private care more quickly, while those unable to afford these services are placed on long waiting lists in the public system. In addition to long waitlists, public PHP services were also reported as not available in several geographic areas, which is in line with the findings by Deslauriers et al. (2017) who found that only 12% of hospitals in Quebec offered PHP services. Another recent study emphasized other disparities in access to PHP services, with lower access observed in rural areas of Ontario, Quebec’s neighboring province (Charette and McLean, 2024). Also concerning from the findings of this study is that even when women were able to pay the high costs or endure long waiting times, the quality of care was reported as inconsistent by some of them. This inconsistency may be due to the fact that there is a great discrepancy between mandatory training requirements to practice PHP in different jurisdictions (Bakker et al., 2018). In addition, physiotherapists may lack access to quality, evidence-based training opportunities (Scordas et al., 2024).

### Strengths and limitations

Strengths of this study includes the use of inductive reflective thematic analysis, which was conducted through a rich iterative process with numerous points of discussion and back-and-forths with the raw data and between team members. This contributed to the rigor and consistency in understanding the participants’ perceptions and experiences. A first limitation is that, although attempts were made to ensure the greatest variability of profiles within the group of participants, there is an underrepresentation of women with lower levels of education, lower socioeconomic backgrounds and those living in remote areas. This may have limited the inclusion of challenges faced by women outside urban settings to access PHP. This may also be due to the fact that participants were recruited through samples of participants in our previous online survey studies. Additionally, as the participants voluntarily agreed to partake in the study, it is possible that they felt more comfortable to discuss pelvic health-related topics, limiting the representation of women who may have had traumatic experiences or for whom pelvic health is a deeply sensitive subject.

In conclusion, the findings from this study offer valuable insights into the perceptions and experiences of women having had PFDs regarding access to PHP services. This work highlights numerous barriers that hinder equitable access to quality PHR services. Our findings point towards the importance for healthcare professionals to advocate for their patients, which includes increasing their knowledge of these conditions and their impacts, which may contribute to avoid the trivialization of PFD symptoms, and ensure that patients are informed about the recommended, evidence-based, treatment options. Healthcare organizations could also play a crucial role in improving access to PHP services, by providing accessible and accurate pelvic health information, and ensuring that quality PHP services are readily available, regardless of factors such as location or financial means.

## Data Availability

All data produced in the present study are available upon reasonable request to the authors.

## ACKNOWLEDGEMENTS

The authors wish to thank all the women who participated in this study. The authors also wish to thank the *Comité stratégique patients-partenaires du Centre Hospitalier Universitaire de Sherbrooke*.

This work was supported by the *Association québécoise de la physiothérapie* and the *Réseau provincial de recherche en adaptation-réadaptation* under a Program 4.6 — Clinical research partnership grant (2020-2021). MM received a salary award from the Fonds de recherche du Québec -Santé.

## REFERENCES

Allaire C, Yong PJ, Bajzak K, Jarrell J, Lemos N, Miller C, Morin M, Nasr-Esfahani M, Singh SS, Chen I 2024 Guideline No. 445: Management of Chronic Pelvic Pain. Journal of Obstetrics and Gynaecology Canada 46: 102283.

Baker L, Phelan S, Snelgrove R, Varpio L, Maggi J, Ng S 2016 Recognizing and Responding to Ethically Important Moments in Qualitative Research. Journal of Graduate Medical Education 8: 607–608.

Bakker E, Shelly B, Esch FH, Frawley H, McClurg D, Meyers P 2018 International Continence Society supported pelvic physiotherapy education guideline. Neurourology and Urodynamics 37: 869–876.

Berre ML, Dumoulin C 2024 Accessibility of Pelvic Floor Physiotherapy for Treating Urinary Incontinence in Older Women in Quebec: An Online Survey. Physiotherapy Canada 76: 86–94.

Boyle R, Hay-Smith EJ, Cody JD, Mørkved S 2014 Pelvic floor muscle training for prevention and treatment of urinary and fecal incontinence in antenatal and postnatal women: a short version Cochrane review. Neurourology and Urodynamics 33: 269–276.

Braun V, Clarke V 2019 Reflecting on reflexive thematic analysis. Qualitative research in sport, exercise and health 11: 589–597.

Busetto L, Wick W, Gumbinger C 2020 How to use and assess qualitative research methods. Neurological Research and Practice 2: 14.

Cardozo L, Rovner E, Wagg A, Wein A, Abrams P 2023 Incontinence 7th Edition. Bristol UK.

Charette M, McLean L 2024 Geographic Accessibility to Pelvic Health Physiotherapy Services Across Ontario: A Geographic Information System Analysis. Physiotherapy Canada. 10.3138/ptc-2023-0114.

Colorafi KJ, Evans B 2016 Qualitative Descriptive Methods in Health Science Research. Health Environments Research & Design 9: 16–25.

Deslauriers S, Raymond M-H, Laliberté M, Lavoie A, Desmeules F, Feldman DE, Perreault K 2017 Access to publicly funded outpatient physiotherapy services in Quebec: waiting lists and management strategies. Disability and Rehabilitation 39: 2648–2656.

Durkin J, Jackson D, Usher K 2020 Qualitative research interviewing: reflections on power, silence and assumptions. Nursing Research 28: 31–35.

Dydyk AM, Gupta N 2024 Chronic Pelvic Pain. In: StatPearls. Treasure Island FL 2024, StatPearls Publishing LLC.

Fante JF, Silva TD, Mateus-Vasconcelos ECL, Ferreira CHJ, Brito LGO 2019 Do Women have Adequate Knowledge about Pelvic Floor Dysfunctions? A Systematic Review. Revista brasileira de ginecologia e obstetricia 41: 508–519.

Gallego-Gómez C, Rodríguez-Gutiérrez E, Torres-Costoso A, Martínez-Vizcaíno V, Martínez-Bustelo S, Quezada-Bascuñán CA, Ferri-Morales A 2024 Urinary incontinence increases risk of postpartum depression: systematic review and meta-analysis. American Journal of Obstetrics & Gynecology 231: 296–307.e211.

Green R, Mardon AK, Beaumont T, Phillips K, Chalmers KJ 2024 The accessibility of pelvic health physiotherapy for adolescents with persistent pelvic pain: a qualitative framework analysis. Physiotherapy Theory and Practice 40: 973–982.

Hawkey A, Chalmers KJ, Micheal S, Diezel H, Armour M 2022 “A day-to-day struggle”: A comparative qualitative study on experiences of women with endometriosis and chronic pelvic pain. Feminism & Psychology 32: 482–500.

Hermansen IL, O’Connell BO, Gaskin CJ 2010 Women’s explanations for urinary incontinence, their management strategies, and their quality of life during the postpartum period. Journal of Wound, Ostomy and Continence Nursing 37: 187–192.

Institut national d’excellence en santé et en services sociaux (INESSS) 2022 Perineal and pelvic rehabilitation for the prevention and treatment of pelvic floor dysfunctions – Part 1: Urinary incontinence.https://www.inesss.qc.ca/fileadmin/doc/INESSS/Rapports/OrganisationsSoins/INESSS_REPP_IU_volet_1_avis.pdf

Jouanny C, Abhyankar P, Maxwell M 2024 A mixed methods systematic literature review of barriers and facilitators to help-seeking among women with stigmatised pelvic health symptoms. BMC Women’s Health 24: 217.

Kepenekci I, Keskinkilic B, Akinsu F, Cakir P, Elhan AH, Erkek AB, Kuzu MA 2011 Prevalence of Pelvic Floor Disorders in the Female Population and the Impact of Age, Mode of Delivery, and Parity. Diseases of the Colon & Rectum 54: 85–94.

Leslie SW, Tran LN, Puckett Y 2024 Urinary Incontinence. In: StatPearls. Treasure Island FL: © 2024, StatPearls Publishing LLC.

Levesque JF, Harris MF, Russell G 2013 Patient-centred access to health care: conceptualising access at the interface of health systems and populations. International Journal for Equity in Health 12: 18.

Melissa D, Lisa MV 2019 Semistructured interviewing in primary care research: a balance of relationship and rigour. Family Medicine and Community Health 7(2): e000057.

Plano Clark VL 2019 Meaningful integration within mixed methods studies: Identifying why, what, when, and how. Contemporary Educational Psychology 57: 106–111.

Scodras S, Yeung E, Colquhoun H, Jaglal SB, Salbach NM 2024 Pelvic Health Content in Canadian Entry-To-Practice Physiotherapy Programs: An Online Survey. Physiotherapy Canada 76: 25–33.

Swahnberg K, Thapar-Björkert S, Berterö C 2007 Nullified: women’s perceptions of being abused in health care. Journal of psychosomatic obstetrics and gynaecology 28: 161–167.

Till SR, As-Sanie S, Schrepf A 2019 Psychology of Chronic Pelvic Pain: Prevalence, Neurobiological Vulnerabilities, and Treatment. Clinical Obstetrics and Gynecology 62: 22–36.

Tunn R, Baessler K, Knüpfer S, Hampel C 2023 Urinary Incontinence and Pelvic Organ Prolapse in Women. Deutsches Ärzteblatt International 120: 71–80.

Zoorob D, Higgins M, Swan K, Cummings J, Dominguez S, Carey E 2017 Barriers to Pelvic Floor Physical Therapy Regarding Treatment of High-Tone Pelvic Floor Dysfunction. Female pelvic medicine & reconstructive surgery 23: 444–448.

